# Vasospasm severity is associated with cerebral infarction after subarachnoid hemorrhage

**DOI:** 10.1101/2021.04.20.21255798

**Authors:** Samuel B Snider, Ibrahim Migdady, Sarah L LaRose, Morgan E Mckeown, Robert W Regenhardt, Pui Man Rosalind Lai, Henrikas Vaitkevicius, Rose Du

**Affiliations:** Division of Neurocritical Care, Department of Neurology, Brigham and Women’s Hospital, Boston MA; Departments of Neurosurgery and Neurology, Massachusetts General Hospital, Boston, MA; Department of Neurosurgery, Brigham and Women’s Hospital, Boston MA; Marinus Pharmaceuticals, Radnor, PA

**Author notes:** **Corresponding Author:** Samuel B. Snider, Division of Neurocritical Care, Department of Neurology, Brigham and Women’s Hospital, 60 Fenwood Road, Boston, MA 02115. Authors contributed equally (Samuel B Snider, Ibrahim Migdady).

## Abstract

**Background:** The presence of angiographic vasospasm after aneurysmal subarachnoid hemorrhage (aSAH) is associated with delayed-cerebral ischemia (DCI)-related cerebral infarction and worsened neurological outcome. Transcranial doppler (TCD) measurements of cerebral blood velocity are commonly used after aSAH to screen for vasospasm. We sought to determine whether time-varying TCD measured vasospasm severity is associated with cerebral infarction and to investigate the performance characteristics of different time/severity cutoffs for predicting cerebral infarction.

**Methods:** We used a retrospective, single-center cohort of consecutive adult aSAH patients with angiographic vasospasm and at least one TCD study. Our primary outcome was DCI-related cerebral infarction, defined as an infarction developing at least 2 days after any surgical intervention without an alternative cause. Time-varying TCD vasospasm severity was defined ordinally (absent, mild, moderate, severe) by the most abnormal vessel on each post-admission hospital day. Cox proportional-hazards models were used to examine associations between time-varying vasospasm severity and infarction. The optimal TCD-based time/severity thresholds for predicting infarction were then identified using the Youden J statistic.

**Results:** Of 218 aSAH patients with angiographic vasospasm, 27 (12%) developed DCI-related infarction. As compared to those without infarction, patients with infarction had higher modified Fisher scale (mFS) scores, and an earlier onset of more-severe vasospasm. Adjusted for mFS, vasospasm severity was associated with infarction (aHR 1.9, 95% CI: 1.3-2.6). A threshold of at least mild vasospasm severity on hospital day 4 had a negative predictive value of 92% for the development of infarction, but a positive predictive value of 25%.

**Conclusions:** In aSAH, TCD-measured vasospasm severity is associated with DCI-related infarction. In a single-center dataset, a TCD-based threshold for predicting infarction had a high negative predictive value, supporting its role as an early screening tool to identify at-risk patients.

## Introduction

Cerebral infarction is an uncommon but devastating complication of delayed cerebral ischemia (DCI) after aneurysmal subarachnoid hemorrhage (aSAH)^1^. Patients with DCI and cerebral infarction following aSAH have particularly low rates of favorable functional outcomes^2^. Previously reported clinical factors associated with infarction after aSAH have included aneurysm size^3^, volume of subarachnoid blood^2, 4, 5^, diabetes^1^, and the presence of an external ventricular drain^1^. The presence of angiographic vasospasm of the large cerebral arteries has the strongest reported association with infarction after aSAH^2, 3, 6^.

Transcranial Doppler (TCD) measurements of blood flow velocities in the large cerebral vessels are routinely used in neurological intensive care units as a non-invasive screening tool to identify patients with vasospasm of the large cerebral arteries^7, 8^. TCD has a high estimated sensitivity (∼90%) but moderate specificity (∼70%) for identifying cerebral vasospasm on conventional angiography^9^. Patients in neurological intensive care units often undergo daily TCD velocity measurements for several weeks following aSAH^10, 11^, requiring a substantial investment in time and resources. In spite of this, time-varying blood flow velocity trends in patients after aSAH have been only reported in small samples of select patients^12^ and have not yet been formally investigated for their ability to predict cerebral infarction. Whether measurable vasospasm precedes the development of infarction and can therefore serve as an alarm in at-risk patients remains unknown.

In this study, we sought to characterize the time varying TCD velocity trends in a retrospective cohort of aSAH patients at high risk for DCI, to determine whether time-varying vasospasm severity is associated with risk of cerebral infarction, and to investigate the performance characteristics of different time/severity cutoffs for predicting cerebral infarction.

## Methods

### Study Cohort

We conducted a retrospective analysis of an IRB-approved, prospectively maintained database of consecutive adult (age > 18) patients with confirmed aneurysmal SAH admitted to an academic medical center between 2011 and 2020 (N = 307). Clinical and demographic information was manually abstracted from the electronic medical record. To enrich our sample for patients with vasospasm, we restricted our analysis to the subset of patients with the mention of ‘vasospasm’ in any CT-angiogram, MR-angiogram, or catheter-angiogram report during their hospital stay (N = 243, Figure 1).

**Figure 1:**
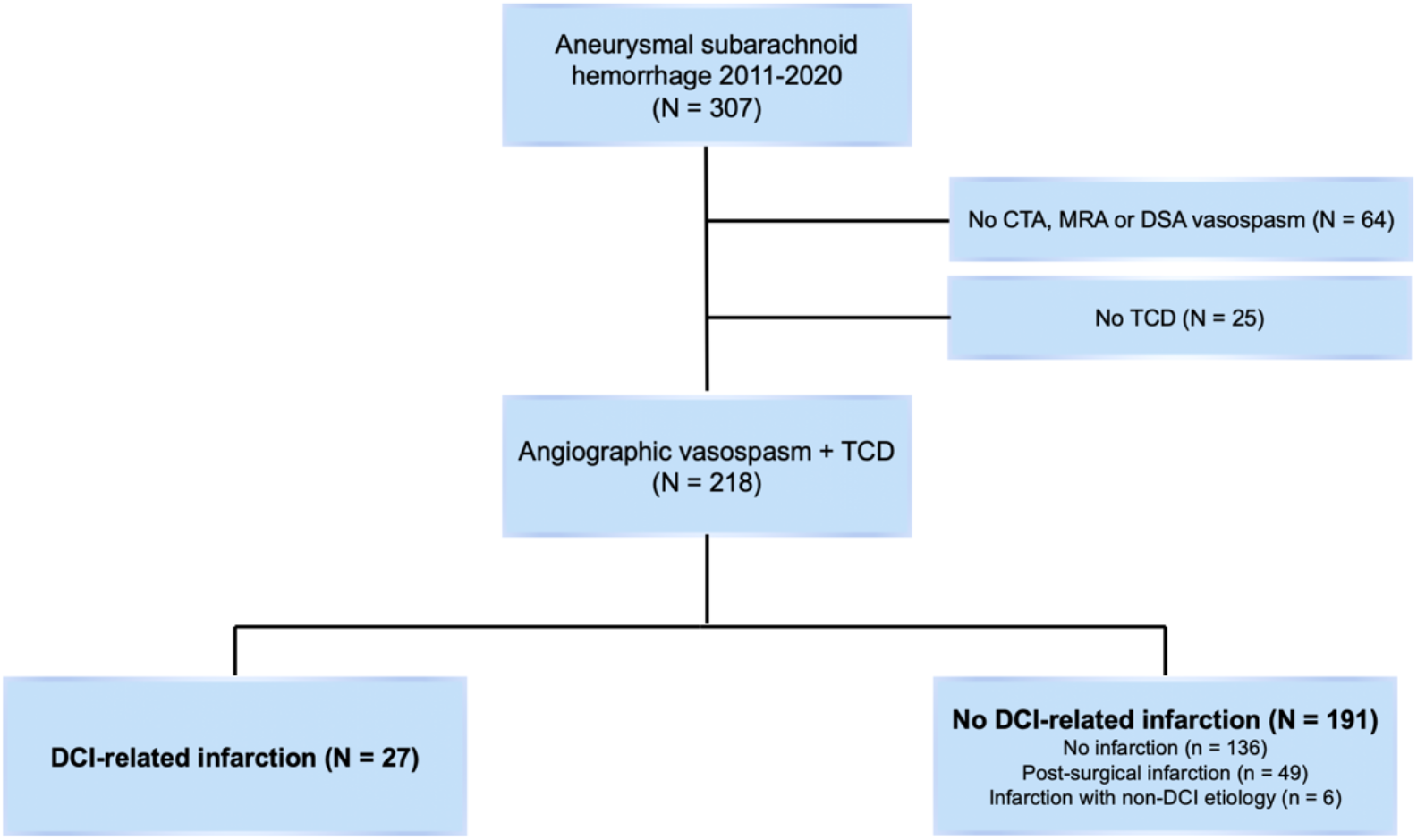
Study Flowchart Abbreviations: CTA: Computed Tomography Angiography, MRA: Magnetic Resonance Angiography, DSA: Digital Subtraction Angiography, TCD: Transcranial Doppler, DCI: Delayed Cerebral Ischemia

### TCD Velocity Measurements

We then cross-referenced this database with a separate, prospectively-maintained database of TCD mean flow velocity (MFV) measurements at this same academic medical center. N = 218 of these patients had at least one TCD study and were included for further analysis (Figure 1). A handheld 2 MHz probe was used to insonate the M1 segment of the middle cerebral artery (MCA), the A1 segment of the anterior cerebral artery (ACA), and the P1 and P2 segments of the poster cerebral artery (PCA) bilaterally. The maximum MFV for each vessel and the date of the recording were stored. Vasospasm severity was categorized based on reported MFV (cm/s) reference ranges for each vessel^13^ as follows: MCA: mild 120 - 149, moderate 150 - 199, severe > 200; ACA: mild 100 - 129, moderate 130 - 149, severe > 150; PCA: mild 80 - 119, moderate 120 - 159, severe > 160.

### Outcome Definition

Our primary outcome was cerebral infarction caused by DCI. We first screened the electronic medical record for infarction by the mention of ‘stroke’, ‘infarction’, or ‘hypodensity’ in radiology reports from any brain CT or MRI obtained during the hospitalization. Each infarction (N = 55) was then adjudicated by two board-certified neurologists (S.B.S. and I.M.). DCI-related infarctions (N = 27) were defined as those that were 1) not present on the admission brain CT, 2) not present within 48 hours of a surgical intervention, and 3) without another clearly identifiable proximate cause (Figure 1). The time of infarction was considered to be the post-admission day on which the infarction was first radiologically documented. The vascular territory of the infarction was defined by manual review of the image on which the infarction was first apparent. Disagreements were resolved by consensus.

### Statistical Analysis

All analyses were performed using RStudio1.1.442 (R Foundation for Statistical Computing, Vienna, Austria; http://www.r-project.org).

### Cohort Comparison

Demographic and clinical characteristics between cohort patients with or without DCI-related infarction were compared using two-sample t-tests (continuous variables) and *χ*^2^ tests (categorical variables).

### Time-Varying Vasospasm Analysis

We defined vasospasm ordinally (absent, mild, moderate, severe) by the most abnormal vessel at the time of each TCD study. Using patient-level vasospasm trajectories beginning on the day of admission (T = 0), a state plot was generated, indicating the proportion of the cohort with each vasospasm severity level at each time point (R: *TraMineR*). Pairwise dissimilarity values for each subject’s vasospasm trajectory were then computed using the ‘optimal matching’ metric with an insertion/deletion cost of 1 and substitution costs defined based on transition rates in the data (R: *TraMineR*). Agglomerative hierarchical clustering of the distance matrix was then conducted using Ward’s method (R: *cluster*). The two most dissimilar clusters were then examined for differences in prevalence of DCI-related infarction.

To determine if time-varying vasospasm severity was associated with infarction, we used a Cox proportional-hazards model (R: *Survival*). Patients were censored either on the day of infarction or on the last day of TCD data obtained during the hospitalization. Our model included time-varying vasospasm severity and admission modified Fisher scale score (mFS), the one covariate found to differ between patients with and without infarction. To determine whether vasospasm severity within a particular vessel was associated with risk of infarction in that vascular territory, we conducted a sensitivity analysis with the most prevalent infarct subtype (right MCA, N = 11). For this sensitivity analysis, we included as exposures time-varying right MCA vasospasm severity either alone or with all of the following covariates: left MCA vasospasm severity, worst ACA vasospasm severity and worst PCA vasospasm severity. The proportional hazards assumption was checked and confirmed for each model (R: *survminer*, all p > 0.15).

To estimate the performance characteristics of static TCD thresholds for predicting subsequent infarction, we computed the sensitivity (Sn), specificity (Sp), positive predictive value (PPV), and negative predictive value (NPV) and plotted receiver operating characteristic (ROC) curves for each level of ordinal vasospasm severity on hospital days 1 through 14. An optimal cut point was chosen based on the Youden’s J statistic.

## Results

### Cohort Characteristics

Among 218 patients with aSAH, radiographic vasospasm, and at least one TCD study, 27 (12%) had DCI-related cerebral infarction (Figure 1, Table 1). Patients with infarction were similar to those without infarction except for having higher mFS scores (Table 1, p = 0.02). The median time between admission and infarction was 8 days (IQR: 6-10, Table 1) and most infarctions (85%) occurred in the anterior circulation (Table 1).

**Table 1:** Cohort Characteristics

| Table 1: Cohort Characteristics |  |  |  |
| --- | --- | --- | --- |
|  | No infarction<br>(n=191) | Infarction<br>(n=27) | P-value |
| Age, mean (95% CI) | 58 (56, 60) | 52 (47, 58) | 0.09 |
| Female, n (%) | 155 (81%) | 21 (78%) | 0.9 |
| White Race | 142 (74%) | 22 (82%) | 0.7 |
| Current Smoker | 72 (38%) | 13 (48%) | 0.4 |
| HTN | 95 (50%) | 11 (41%) | 0.5 |
| Aneurysm location |  |  | 0.3 |
| MCA | 34 (18%) | 3 (11%) |  |
| ACA | 64 (34%) | 14 (52%) |  |
| ICA | 66 (35%) | 8 (30%) |  |
| Posterior | 27 (14%) | 2 (7%) |  |
| Hunt & Hess Grade |  |  | 0.06 |
| 1 | 37 (19%) | 2 (7%) |  |
| 2 | 54 (28%) | 3 (11%) |  |
| 3 | 44 (23%) | 9 (33%) |  |
| 4 | 33 (17%) | 9 (33%) |  |
| 5 | 22 (12%) | 4 (15%) |  |
| Modified Fisher Scale |  |  | 0.02 |
| 1 | 10 (5%) | 0 (0%) |  |
| 2 | 15 (8%) | 1 (4%) |  |
| 3 | 79 (41%) | 5 (19%) |  |
| 4 | 86 (45%) | 21 (78%) |  |
| Treatment with Clip | 113 (59%) | 17 (63%) | 0.9 |
| Time to Infarction (days),<br>median (IQR) | - | 8 (6, 10) | -- |
| Infarct Vascular Territory |  |  | -- |
| MCA | - | 14 (52%) |  |
| ACA | - | 9 (33%) |  |
| PCA | - | 5 (19%) |  |
| Basilar |  | 1 (4%) |  |

### TCD-measured Vasospasm Severity Trends

State plots show a broad peak in the incidence of TCD-measured vasospasm between 7 and 21 days (Figure 2A). The two most disparate clusters of TCD velocity trajectories were a group with mild, delayed spasm (Cluster 1, Figure 2B) and a group with earlier, severe spasm (Cluster 2, Figure 2B). 8% of Cluster 1 patients had an infarction compared to 20% of Cluster 2 patients (*χ*^2^ _(1,218)_ = 5.4, p = 0.02). Patients with infarction developed TCD-measured vasospasm earlier (*with:* median 3 days, IQR: [2, 5], *without:* 4 [3, 6], p = 0.03) and reached peak spasm earlier (*with*: 6 days [4, 8], *without:* 7 [5, 11], p = 0.04) than those without infarction.

**Figure 2:**
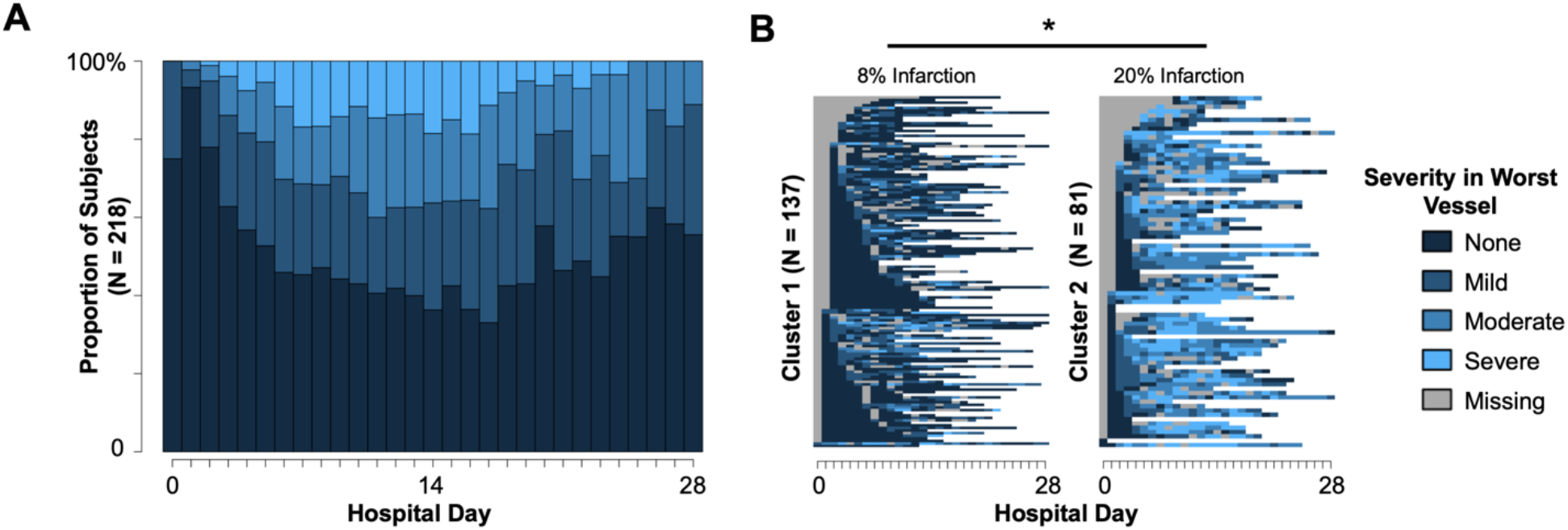
Severity of TCD-measured vasospasm after SAH (A) State plot of the proportion of the cohort with each ordinal level of vasospasm severity (absent, mild, moderate, severe) between day of admission (0) and hospital day 28. (B) Individual patient trajectories from the two most-dissimilar clusters of vasospasm trajectories. Missing is shaded grey. Rates of infarction significantly differed between clusters.

### TCD-measured Vasospasm as a Predictor of Infarction

Controlling for the mFS, the time-varying vasospasm severity was associated with the development of infarction [adjusted Hazard Ratio (aHR) 1.9 per unit increase in spasm severity, 95% CI: 1.3 - 2.6, p = 0.0004, unadjusted HR 2.0, (1.4, 2.8), Figure 3]. To determine if the severity of vasospasm in a particular vessel was associated with the risk of infarction in that vascular territory, we conducted a sensitivity analysis using the most prevalent subgroup of infarctions: those occurring within the right MCA territory (N

**Figure 3:**
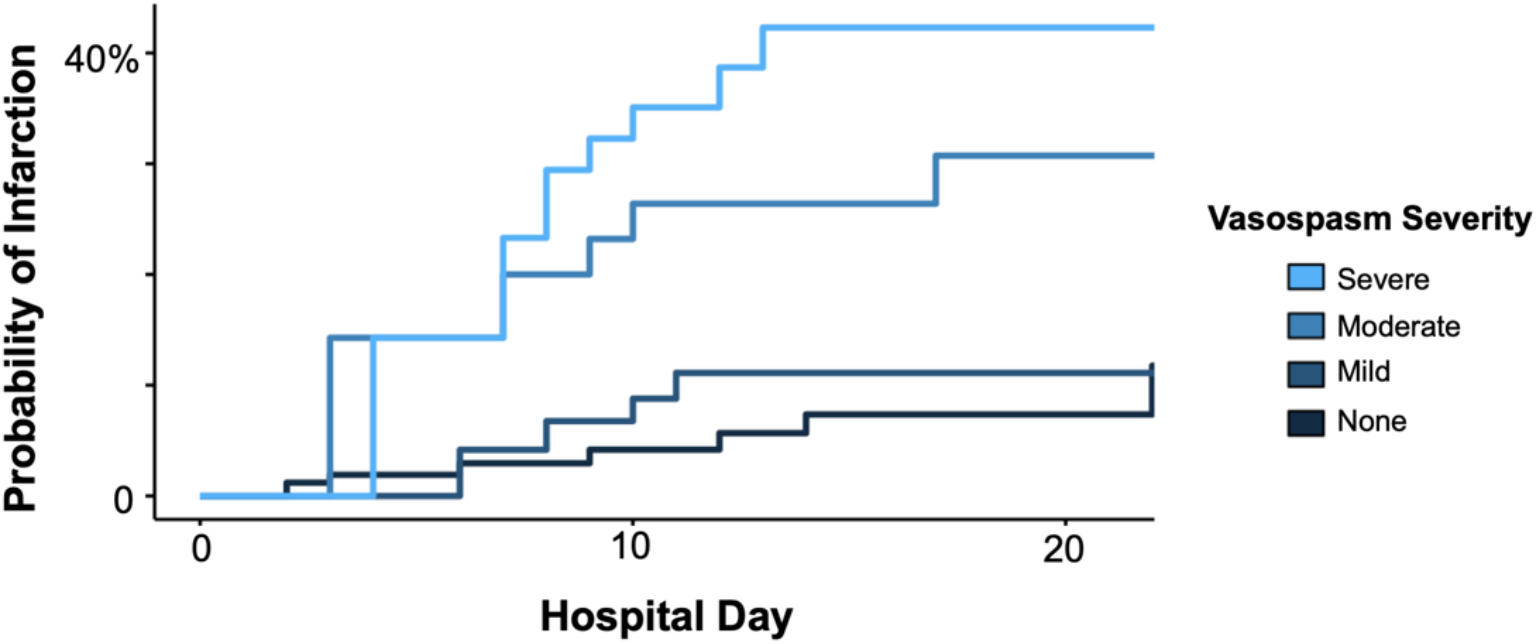
Risk of infarction increases with vasospasm severity Kaplan-Meier curves for the probability of infarction by time-varying vasospasm severity.

= 11). Vasospasm severity within the right MCA was associated with right MCA territory infarction (HR 2.5, [1.5, 4.2], p = 0.0005), and trended towards significance even after controlling for the vasospasm severity in all other vascular territories (aHR 1.8, [1.0, 3.4] p = 0.05).

Finally, we sought to evaluate the infarct-prediction performance of TCD-measured vasospasm severity thresholds at specific time points during the hospitalization. Optimal discrimination was achieved using a threshold of at least mild vasospasm severity on hospital day 4 (Figure 4A, *red circle*, Sn 59%, Sp 67%), though the PPV was only 25%. This time/severity threshold also had the highest NPV (92%) for ruling out the subsequent development of infarction. Any thresholds with a PPV greater than 25% occurred at sensitivities under 25% (Figure 4B).

**Figure 4:**
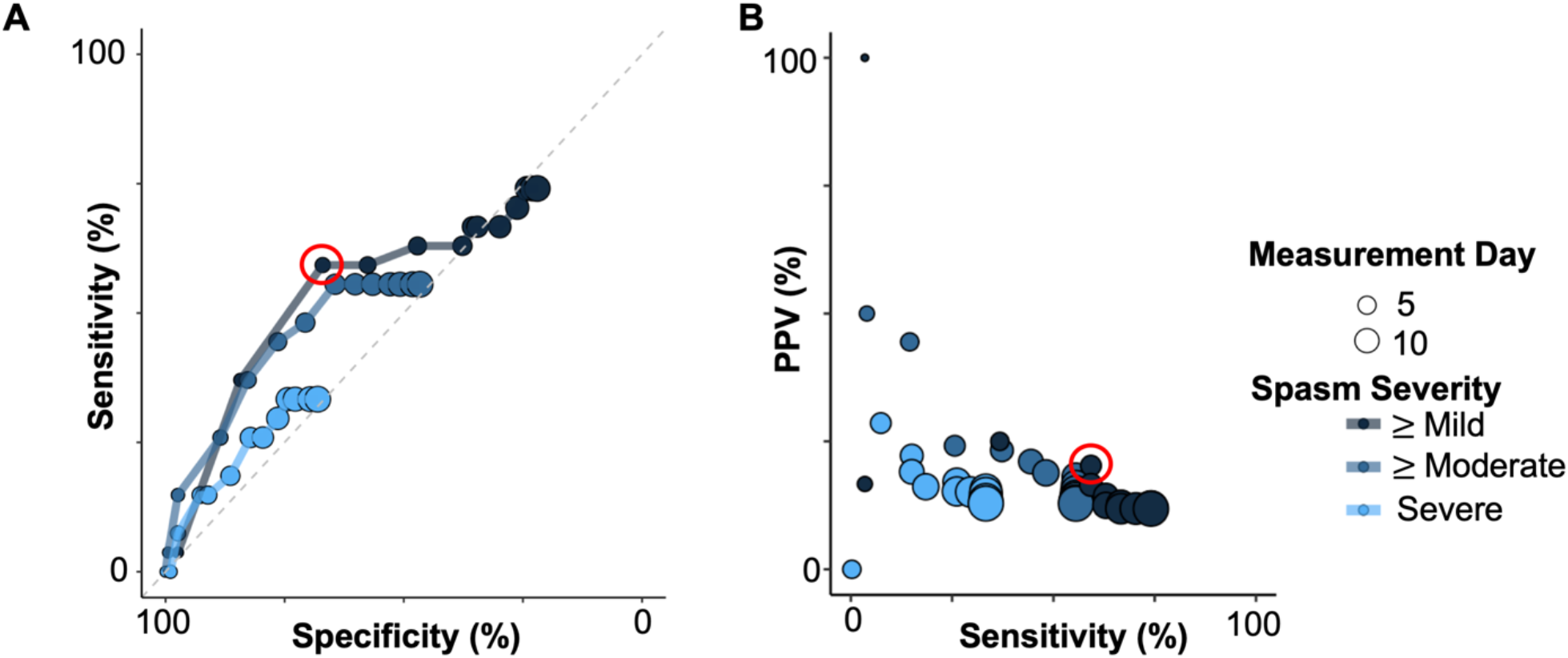
Performance characteristics of different time/severity thresholds (A) Receiver Operator Characteristic (ROC) plots for varying vasospasm severity (color) and post-admission day (circle size) thresholds for predicting infarction. The Youden-J statistic is maximized at a threshold of at least mild severity on hospital day 4 (red circle). (B) The positive predictive value (PPV) of each threshold is plotted against its sensitivity, with the optimal discriminating point from (A) indicated within the red circle.

## Discussion

We evaluated TCD-based cerebral blood flow velocity trends in a large cohort of aSAH patients, identifying an association between vasospasm severity and DCI-related cerebral infarction. We confirmed previous reports that vasospasm incidence rises and peaks between 7 and 21 days after the initial hemorrhage. Patients with infarction were enriched in a data-driven cluster of vasospasm trajectories with earlier vasospasm onset times and shorter times to peak vasospasm severity. Controlling for clinical SAH grade, TCD-measured vasospasm severity was associated with risk of infarction. The severity of spasm in a given vessel was associated with risk of infarction in the territory supplied by that vessel. Finally, we characterized a range of TCD time and severity thresholds for predicting infarction, finding that the presence of at least mild vasospasm on hospital day 4 has the best discriminative value.

### Vasospasm Time-Course

Our report aligns well with descriptions of vasospasm time course derived from classic angiographic studies of vessel diameter in aSAH patients^14^. Our work benefits from a higher sampling rate (daily measurements) for deriving patient-level trajectories than is possible with conventional angiography. While an analysis of angiography data has suggested that vasospasm does not extend beyond 12 days after aSAH^14^, we find that elevated cerebral blood flow velocities persist longer than that.

### Association with Infarction and Predictive Characteristics

Higher TCD-measured blood flow velocities are associated with higher rates of DCI-related infarction. Furthermore, blood flow velocities within a particular vessel are associated with risk of infarction in the territory supplied by that vessel, even controlling for vasospasm severity in other vessels. While there is active debate about whether large vessel vasospasm is a cause or epiphenomenon of DCI^15^, the spatial specificity of these findings support a causal role.

Consistent with the known limitations of using TCD-measured elevated cerebral blood flow velocities to identify angiographic vasospasm^9, 16^, the optimal predictors of infarction had much higher NPVs than PPVs. Flagging patients who reach at least mildly severe vasospasm in any vessel by hospital day 4 would result in a low rate of missed infarctions (8%), though only one in four of these patients would ultimately go on to develop an infarction. This time/severity threshold could therefore be clinically useful as a trigger to reduce the frequency of monitoring.

### Limitations

DCI has been shown to associate with poor outcome after aSAH independently of cerebral infarction^2^. However, DCI is difficult to define objectively, particularly in a retrospective cohort. While patients with infarction may only represent a subset of those with DCI, they likely represent the most severe subset.

Given that we did not measure or account for spasmolytic treatments, the vasospasm severities reported here should therefore be interpreted as a lower bound for the untreated severity. We would expect this limitation to bias us towards the null. However, if elevated TCD-measured blood flow velocities triggered an increased frequency of interventions that can themselves cause infarction (iatrogenic blood pressure augmentation, catheter angiograms), then a confound could be driving the positive association. Furthermore, while we took precautions to avoid including infarctions that were likely a consequence of the surgical treatment of the aneurysm, there were certain cases occurring within 4 days of surgery where a post-surgical infarct was impossible to exclude. However, this limitation should only reduce our power to identify significant associations.

All time-varying analysis treated the admission day as the day of aneurysmal rupture. If patients ruptured days before presenting to the hospital, their measured vasospasm onset times would have been systematically earlier (relative to hospital admission day). We would expect the probability of this occurring to be higher in patients with lower-mFS hemorrhages, and thus higher in patients without DCI-related infarction. This systematic error in measuring the post-bleed time would thus be more likely to bias us against significant associations and towards worse predictive performance. We did not record the presence of focal neurological deficits, the indication for obtaining brain imaging, or the frequency of imaging obtained during the hospitalization. If patients with DCI-related infarctions were more likely to have undergone MRI without being more likely to have experienced a typical indication for scanning, then we may have missed a certain proportion of DCI-related infarctions. However, this would only diminish the significance of any measured findings.

Finally, these findings reflect a retrospective analysis of a single-center cohort, and thus may have limited generalizability unless externally validated.

## Conclusions

We provide the first systematic analysis of TCD velocity trends in a large, retrospective dataset of aSAH patients. We demonstrate that vasospasm severity is associated with the development of DCI-related cerebral infarction in a vessel-specific manner. Finally, we show that a threshold of at least mild vasospasm severity on hospital day 4 has the best performance characteristics for identifying patients at risk for developing cerebral infarction.

## Data Availability

De-identified data available on reasonable request to the corresponding author.

## Acknowledgements

none

